# Mechanisms of change in a dog-assisted psycho-oncological group intervention: An explorative qualitative interview study

**DOI:** 10.64898/2025.12.16.25342361

**Authors:** Cheyenne Topf, Mareike Rutenkröger, Isabelle Scholl, Pola Hahlweg

## Abstract

**Background:** Cancer survivors face substantial psychosocial stress in addition to disease-related challenges, often resulting in mood and anxiety disorders. Psycho-oncological interventions, such as group psychotherapy, are key in addressing these issues. Dog-assisted interventions also have the potential to alleviate psychosocial stress. This study aimed at investigating experienced mechanisms of change of a dog-assisted psycho-oncological group intervention (DAPOGI).

**Methods:** An exploratory cross-sectional qualitative study with semi-structured interviews with group participants (i.e., participating cancer survivors) and HCPs, (i.e., psychologists, dog handler) of a DAPOGI for highly anxious cancer survivors was carried out. Data was analyzed using qualitative content analysis.

**Results:** In eleven interviews with eight group participants and three HCPs, various mechanisms of change were identified, particularly related to the group setting (e.g., sharing experiences for emotional release) and the presence of the dog (e.g., calming effects, promoting cohesion and therapeutic relationships). Various mechanisms were reported, emphasizing the dog’s positive impact on the overall group therapy experience. We found great overlap between participants’ and HCPs’ experiences.

**Discussion and conclusion:** This study suggests that DAPOGIs may be a valuable way to support psychologically distressed cancer survivors by enhancing traditional psycho-oncological group therapies through unique dog-related mechanisms of change. These mechanisms appear to surpass those of general group psychotherapy, fostering more positive experiences for group participants and HCPs alike. While these findings are encouraging, further research and clinical trials are needed to confirm the effectiveness of DAPOGIs and refine their clinical application.

## BACKGROUND

Cancer survivors (i.e., people living with or after a cancer diagnosis) face substantial psychosocial burden [1]. Hence, they were found to be more at risk to develop a mental disorder compared to the general population [2,3]. In a recent meta-analysis involving 135,015 cancer survivors in over 30 countries, the prevalence was 23.7% for depression and 24.4% for anxiety disorders [4]. Furthermore, a systematic review showed that fifty-eight percent of cancer survivors indicated fear of cancer recurrence and progression [5].

Group therapy is an established form of psychotherapy to alleviate psychosocial burden in cancer survivors [6,7] and is recommended in clinical guidelines [8]. Group therapy for cancer survivors has demonstrated moderate effects on depressive and anxiety symptoms and small effects on psychosocial distress and health-related quality of life [9]. For group psychotherapy in general, Yalom and Leszcz [10] proposed eleven mechanisms of change (i.e., factors that facilitate the psychotherapeutic process and contribute to therapeutic changes in patients, [11]). These include for example the *instillation of hope* through seeing that change is possible when others get better, or the experience of *universality* (i.e., the sense of not being the only person). Sherman et al. [12] further delineated various treatment effects in group interventions for cancer survivors, including heightened social support, increased illness understanding, facilitated emotional disclosure, and enhanced existential meaning. The therapeutic benefits of structured cognitive-behavioral groups are likely tied to skill development in coping and self-management, while less structured, interactive group formats may engender profound existential shifts or deeper emotional processing [12].

In addition to established psycho-oncology interventions, animal-assisted interventions (AAIs) hold promise for cancer survivors by using animals to positively influence their experience and behavior [13]. Dog-assisted therapy (DAT), as a specific form of AAIs, is emerging as a viable approach to alleviating psychosocial distress by utilizing the calming and mood-enhancing effects of dogs [14–17]. Research suggests reduced anxiety, improved social and coping skills, increased self-efficacy, and enhanced positive affect [14,15]. Shen et al. identified several key mechanisms of effectiveness for AAIs in group settings, including fostering feelings of normalcy, improving behavioral activation, physical contact, calming and comforting, and distraction [18]. Despite studies investigating pet visits during radiation and chemotherapy for cancer survivors [19,20], research on AAI or DAT in outpatient group settings remains limited [14,15].

Considering this research gap, we combined both interventions in a dog-assisted psycho-oncological group therapy for highly anxious cancer survivors. Combining the mechanisms of change from both interventions holds potential to enhance the effects of psycho-oncological group therapy. Hence, this study aimed to investigate how the participating cancer survivors, clinical psychologists, and dog handler experienced the mechanisms of change in this dog-assisted psycho-oncological group intervention (DAPOGI) for highly anxious cancer survivors.

## METHODS

### Study design and group therapy intervention

We conducted exploratory qualitative interviews with cancer survivors (hereafter referred to as group members, GM), clinical psychologists, and the therapy dog handler (hereafter jointly referred to as healthcare professionals, HCPs) who had participated in the DAPOGI for highly anxious cancer survivors. We adhered to the Consolidated Criteria for Reporting Qualitative Research (COREQ) guidelines (cp. Supplementary Material 1[21]).

The DAPOGI was conducted within routine care, with the scientific evaluation performed subsequently. It had run for seven 90-minute sessions from January to September 2020 at the outpatient clinic for psycho-oncology of the University Medical Center Hamburg-Eppendorf. Due to the COVID-19 pandemic, it had to be paused between March and August. Adult cancer survivors experiencing at least moderate psychological distress and anxiety were eligible for participation (NCCN Distress Thermometer > 6 [22], GAD-7 > 10 [23]). Experienced clinical psychologists, trained in cognitive-behavioral therapy, accompanied by a certified therapy dog and its handler led the DAPOGI sessions. Each session followed a clear structure (beginning, working phase, end). The intervention aimed to foster anxiety management and self-compassion through an integrative approach, including cognitive-behavioral, existential, trauma, and compassion-based elements. Psychoeducational components, relaxation, mindfulness exercises, and the therapy dog as a group mediator and emotional support were supposed to enrich the therapeutic experience.

### Materials and questionnaires

To ensure consistency, structured interview guides were developed for group members and HCPs (cp. Supplementary Material 2a and 2b) and covered the following topics: prior experiences with dogs and/or group therapy, motivation and expectations for the DAPOGI, experiences during DAPOGI sessions and with the therapy dog. To describe the sample, group members completed a demographic questionnaire, HCPs were asked about their professional experience during the interview (cp. Supplementary Material 3).

### Recruitment and data collection

All group members and HCPs who participated in the DAPOGI were invited to this study (purposive sampling). The principal investigator and senior author of this study (PH) was also one of the clinical psychologists leading the DAPOGI, thus in a double role. PH was interviewed as her additional perspective was deemed more valuable in this exploratory study than the benefits of restricting her to one role. Safeguards against undue influence of her double role are described below.

PH reached out to group members, the second clinical psychologist, and the dog handler about participation. Due to the double-role of PH, all subsequent steps of informing about the study, obtaining informed consent, and conducting the interview were undertaken by the first author (CT) without PHs involvement. If participants voiced interest, CT, a female master’s student with experience in psychological research and interviewing, who had not been involved in the DAPOGI, contacted them by phone and informed them about the study’s objectives and procedures. In case of continued willingness to participate, an interview appointment was scheduled, group members received written informed consent forms by mail, and had to provide written informed consent before the interview. The one-time semi-structured individual interviews were conducted via telephone by CT and audio-recorded. No field notes were taken. Participants did not receive compensation. Transcripts and results were not returned to participants for feed-back.

### Data analysis

Interview recordings were transcribed verbatim and anonymized. Data analysis followed Mayring’s and June’s [24] qualitative content analysis approach, conducted inductively, focusing on statements where participants articulated mechanisms of change of the DAPOGI. To safeguard data analysis, the following three perspectives were included: CT, who had conducted the interviews, but was not involved in the DAPOGI; PH, who had participated in the DAPOGI and given an interview, but did not conduct interviews; MR, who had been involved in neither the DAPOGI nor conducting the interviews. Initial coding was conducted independently for group members and HCPs due to their difference in perspectives. CT coded half of the data set. PH provided feedback on the preliminary codings and the coding system and CT and PH discussed it. CT incorporated agreed-upon changes before analyzing remaining data. CT then summarized the codes axially. PH and MR provided feedback on the pre-final and hierarchically structured coding system and CT revised the coding system after joint discussion. Due to substantial overlap, coding systems of group members and HCPs were merged at this point. Transcription used f4transkript (version 4.2, dr. dresing & pehl, GmbH, Marburg), data analysis was facilitated by MAXQDA [25]. Descriptive statistics were computed using SPSS [26].

## RESULTS

### Sample characteristics

Eleven interviews were conducted with eight group members, two clinical psychologists, and one dog handler. Duration ranged from 20-60 minutes (mean=41.51, *SD*=15.32). Group members’ mean age was 56 years (*SD*=8.9 years, range from 44 to 75 years) and 75% were female. Approximately two-thirds indicated a high level of formal education (>11 years at school; *n*=5, 62.5%). Predominant tumor entities were breast cancer (*n*=3, 37.5%) and gynecological cancers (*n*=2, 25%). 62.5% had been diagnosed within the last 5 years. Cancer was localized in approximately one third (n=3, 37.5%), metastasized or in remission in one quarter each (n=2, 25%). 62.5% had never lived with a dog. Group members reported diverse experiences with psychosocial support. Most had undergone individual psychotherapy (*n*=6, 75%) and half had participated in group psychotherapy (*n*=4, 50%) in the past. None had prior AAI experience. Additional demographic details are provided in Table 1.

**Table 1.**
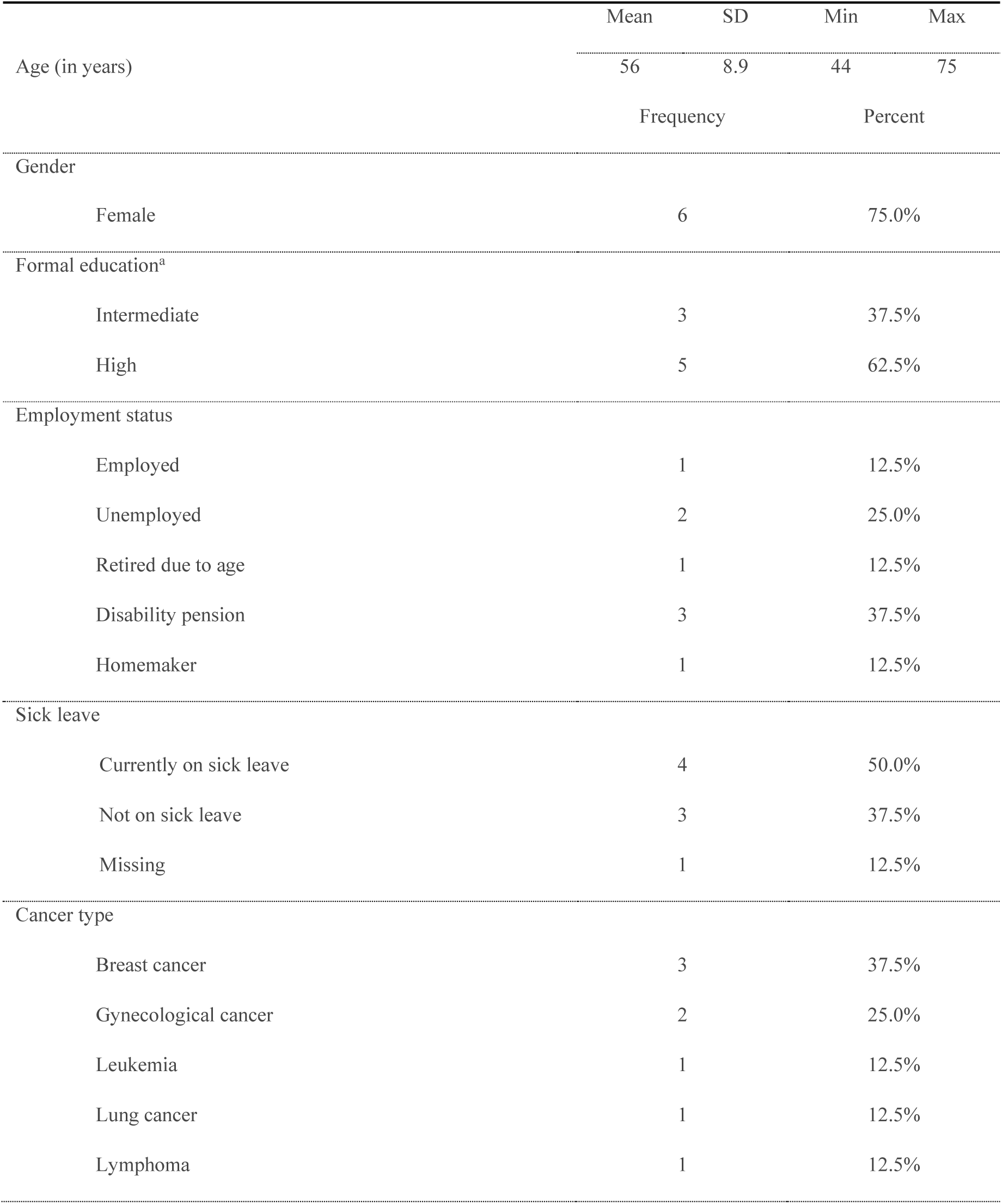
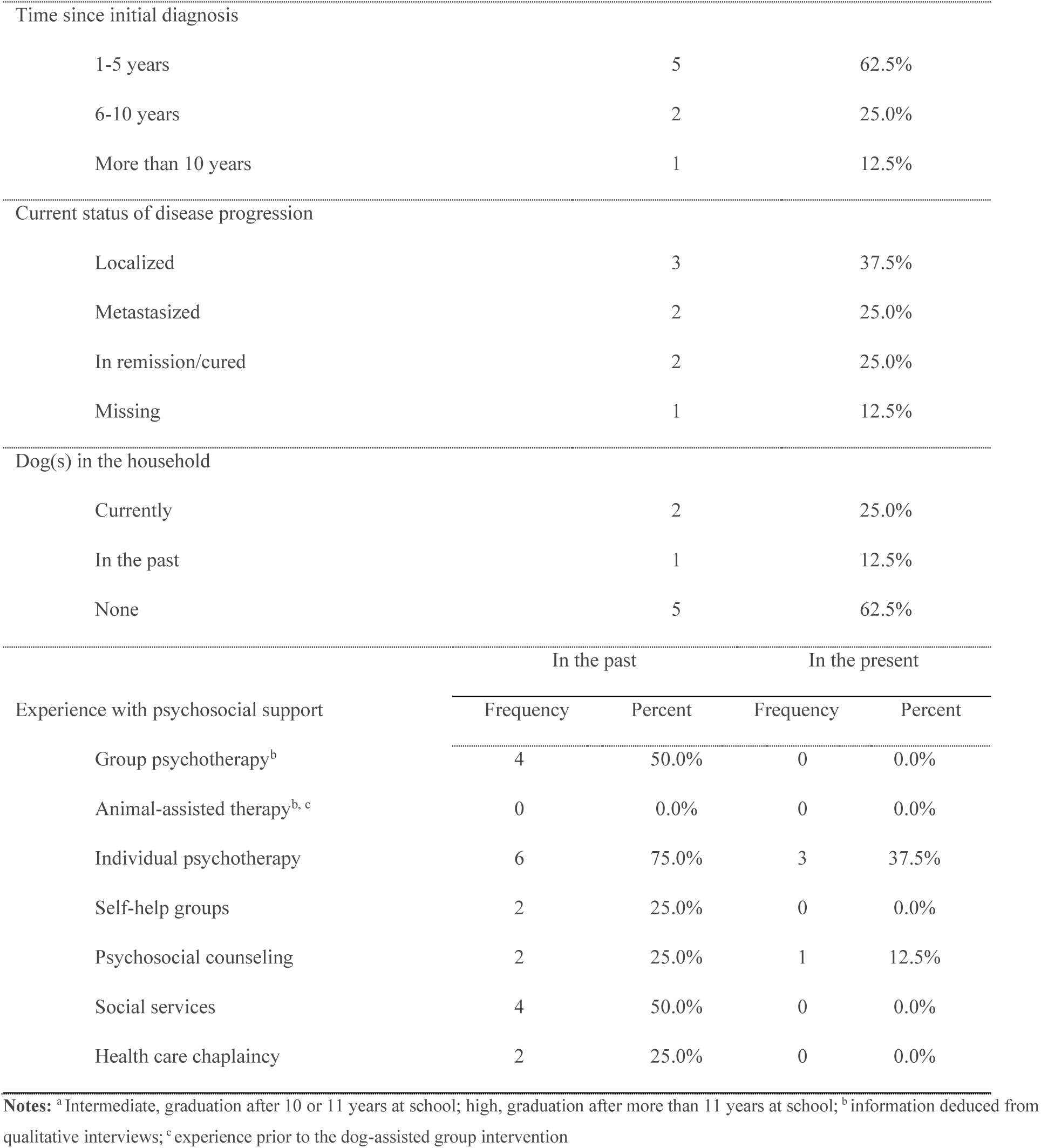
Descriptive characteristics of the group members (n=8).

One clinical psychologist had six, the other ten years of experience in psycho-oncology. The dog handler had seven years of experience working with therapy and service dogs. All had prior group therapy experiences and one clinical psychologist and the dog handler had been involved in a previous iteration of the DAPOGI. All of them had experiences with family dogs’ and both clinical psychologists had prior encounters with therapy animals.

### Mechanisms of change

This section elaborates the mechanisms of change reported by interview participants, organized into four domains (Figure 1). The corresponding coding system with descriptions and quotes for each identified mechanism of change is provided in Supplementary Material 4.

**Figure 1.**
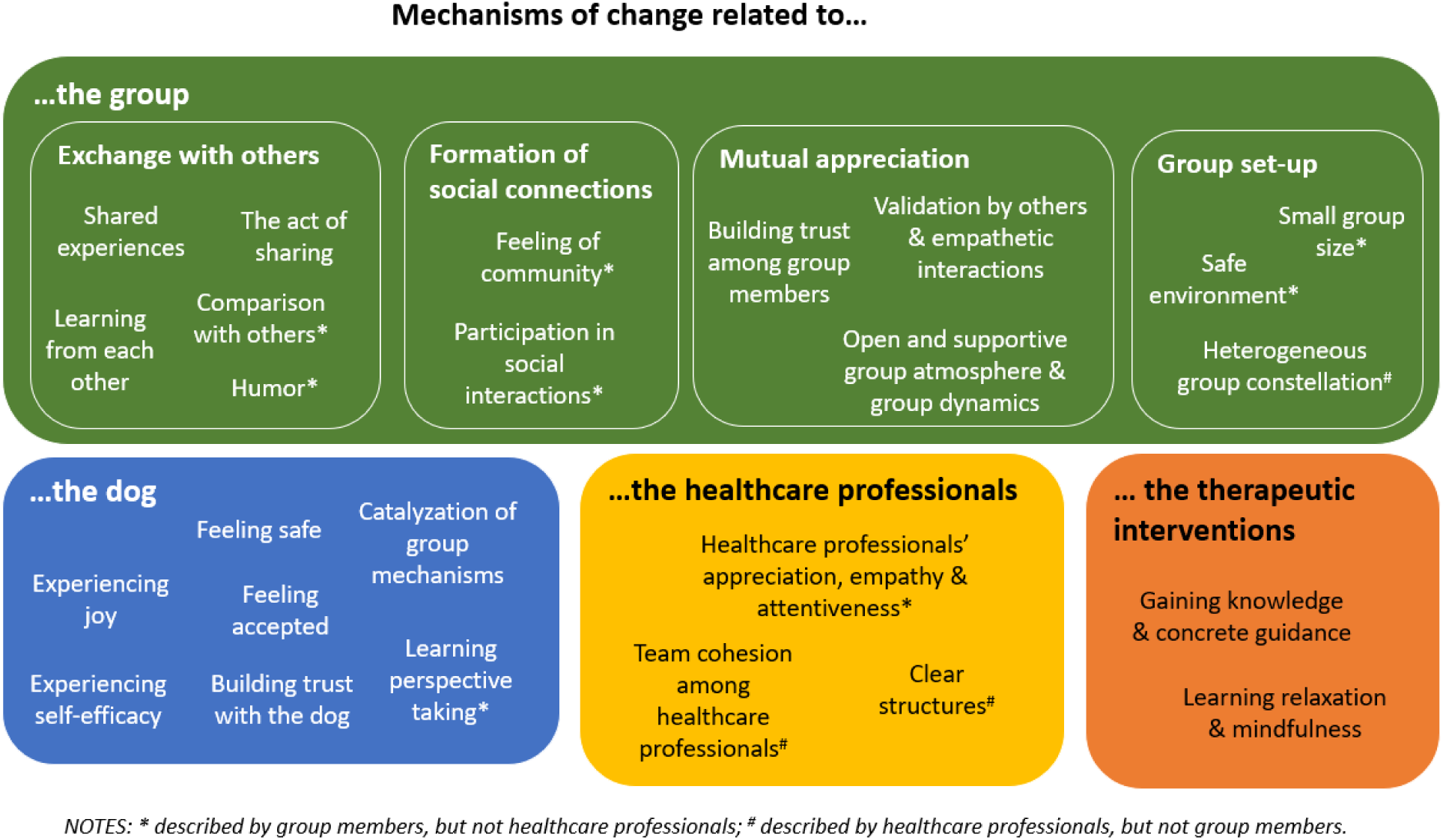
Mechanisms of change reported by group members and HCPs of the DAPOGI for highly anxious cancer survivors.

#### Mechanisms of change related to the group

We sorted the mechanisms of change experienced at group level into four sub-domains. Illustrative quotes for the mechanisms of change related to the group are provided in Table 2.

**Table 2.**
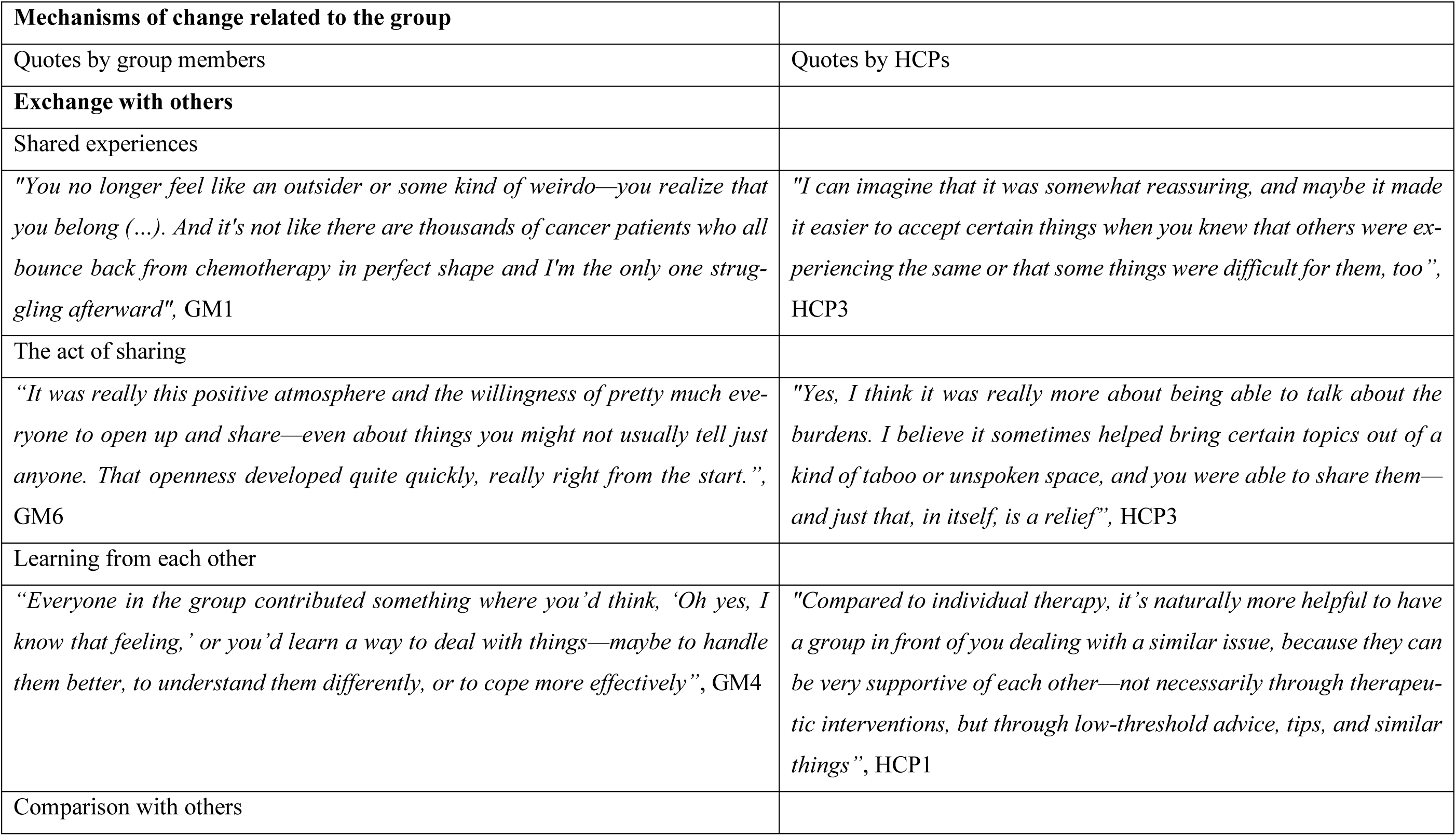
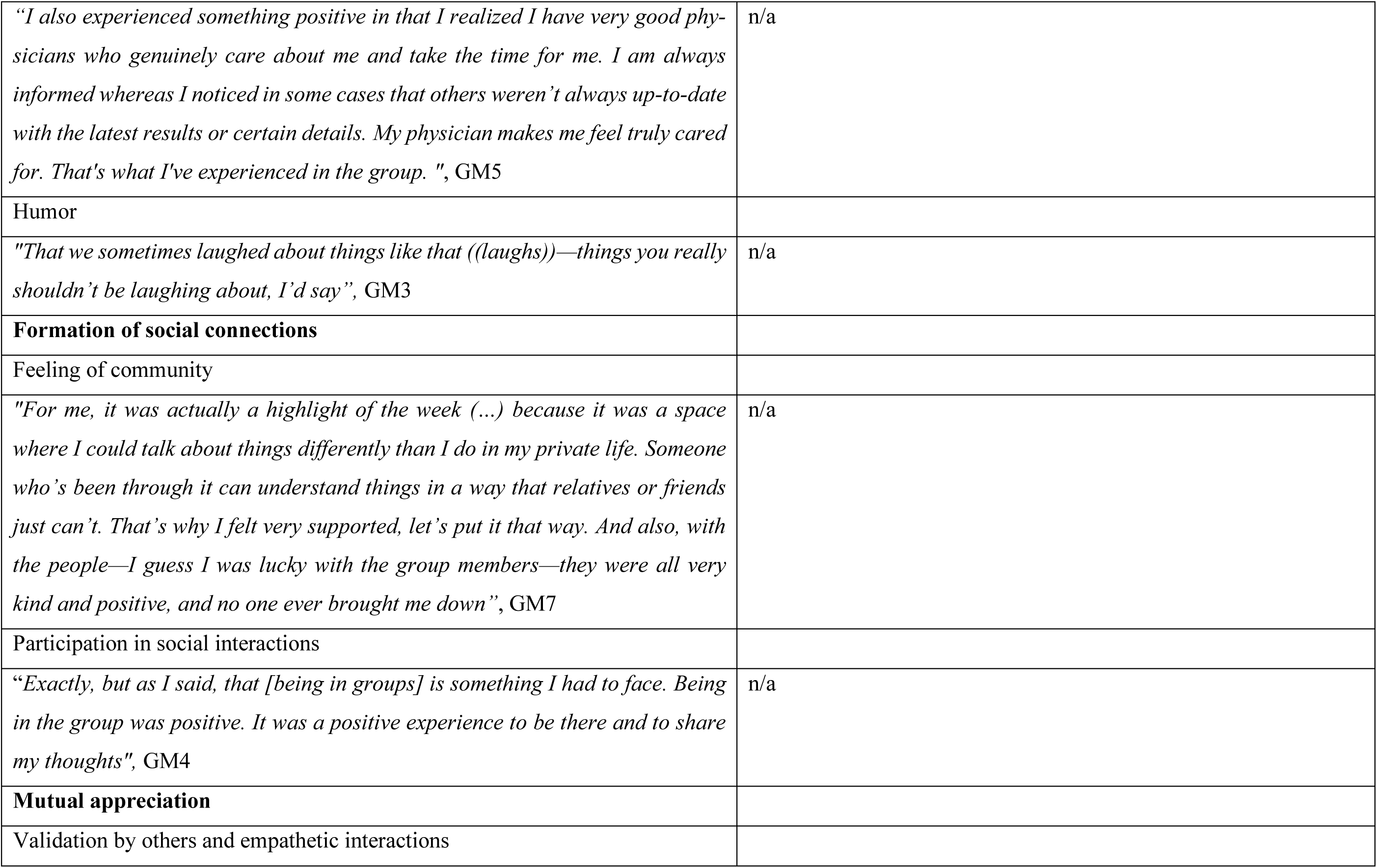
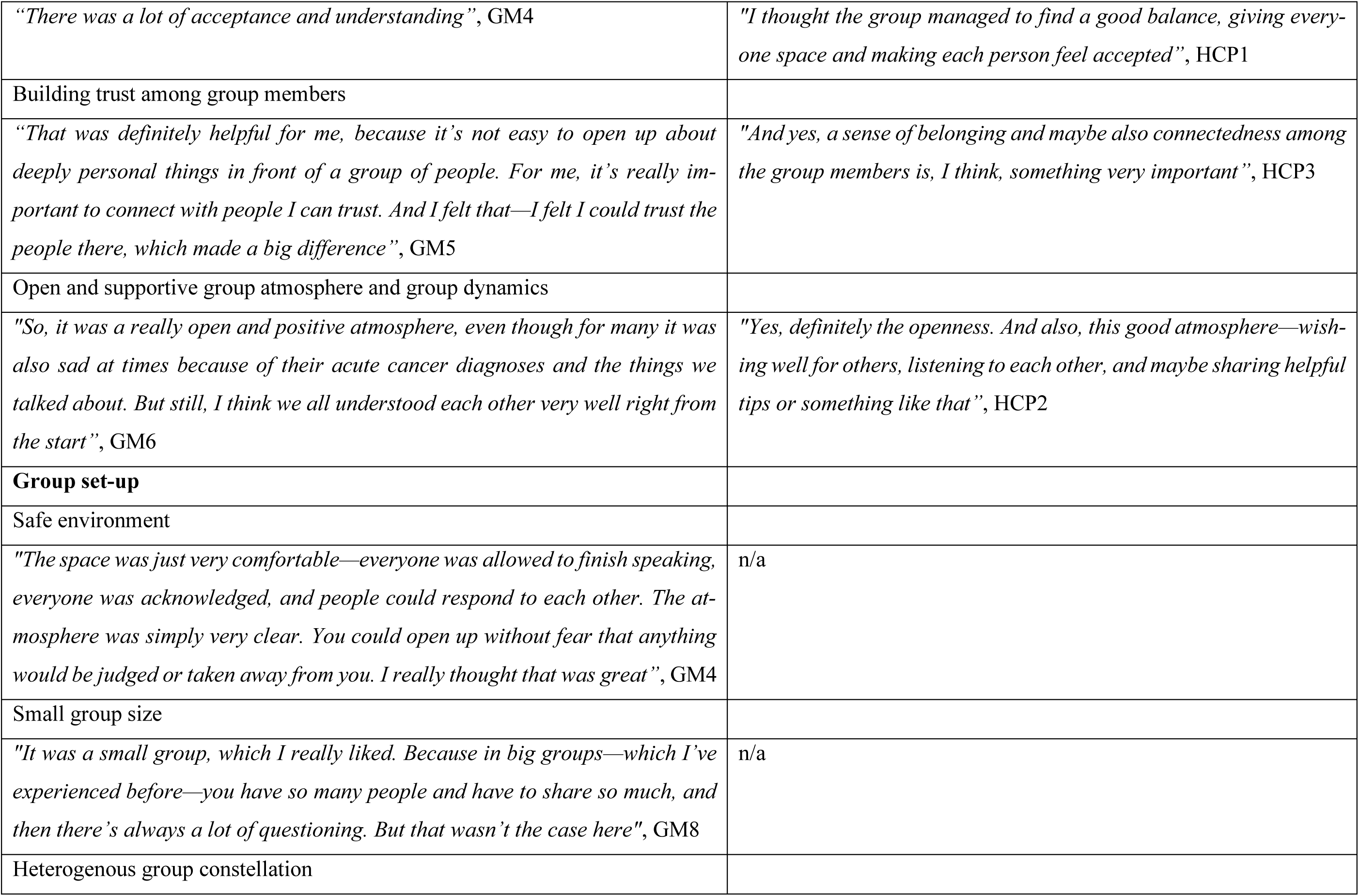
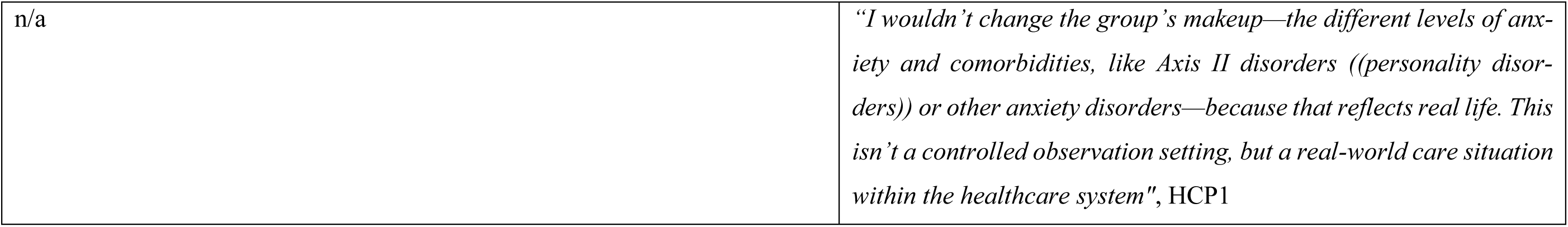
Illustrative quotes by the domain Mechanisms of change related to the group.

|  |  |
| --- | --- |
| **Mechanisms of change related to the group** |  |
| Quotes by group members | Quotes by HCPs |
| **Exchange with others** |  |
| Shared experiences |  |
| *"You no longer feel like an outsider or some kind of weirdo—you realize that you belong (...). And it's not like there are thousands of cancer patients who all bounce back from chemotherapy in perfect shape and I'm the only one struggling afterward", GM1* | *"I can imagine that it was somewhat reassuring, and maybe it made it easier to accept certain things when you knew that others were experiencing the same or that some things were difficult for them, too", HCP3* |
| The act of sharing |  |
| *"It was really this positive atmosphere and the willingness of pretty much everyone to open up and share—even about things you might not usually tell just anyone. That openness developed quite quickly, really right from the start.", GM6* | *"Yes, I think it was really more about being able to talk about the burdens. I believe it sometimes helped bring certain topics out of a kind of taboo or unspoken space, and you were able to share them—and just that, in itself, is a relief", HCP3* |
| Learning from each other |  |
| *"Everyone in the group contributed something where you'd think, 'Oh yes, I know that feeling,' or you'd learn a way to deal with things—maybe to handle them better, to understand them differently, or to cope more effectively", GM4* | *"Compared to individual therapy, it's naturally more helpful to have a group in front of you dealing with a similar issue, because they can be very supportive of each other—not necessarily through therapeutic interventions, but through low-threshold advice, tips, and similar things", HCP1* |
| Comparison with others |  |
| *"I also experienced something positive in that I realized I have very good physicians who genuinely care about me and take the time for me. I am always informed whereas I noticed in some cases that others weren't always up-to-date with the latest results or certain details. My physician makes me feel truly cared for. That's what I've experienced in the group. ", GM5* | n/a |
| Humor |  |
| *"That we sometimes laughed about things like that ((laughs))—things you really shouldn't be laughing about, I'd say", GM3* | n/a |
| **Formation of social connections** |  |
| Feeling of community |  |
| *"For me, it was actually a highlight of the week (...) because it was a space where I could talk about things differently than I do in my private life. Someone who's been through it can understand things in a way that relatives or friends just can't. That's why I felt very supported, let's put it that way. And also, with the people—I guess I was lucky with the group members—they were all very kind and positive, and no one ever brought me down", GM7* | n/a |
| Participation in social interactions |  |
| *"Exactly, but as I said, that [being in groups] is something I had to face. Being in the group was positive. It was a positive experience to be there and to share my thoughts", GM4* | n/a |
| **Mutual appreciation** |  |
| Validation by others and empathetic interactions |  |
| *"There was a lot of acceptance and understanding", GM4* | *"I thought the group managed to find a good balance, giving everyone space and making each person feel accepted", HCP1* |
| Building trust among group members |  |
| *"That was definitely helpful for me, because it's not easy to open up about deeply personal things in front of a group of people. For me, it's really important to connect with people I can trust. And I felt that—I felt I could trust the people there, which made a big difference", GM5* | *"And yes, a sense of belonging and maybe also connectedness among the group members is, I think, something very important", HCP3* |
| Open and supportive group atmosphere and group dynamics |  |
| *"So, it was a really open and positive atmosphere, even though for many it was also sad at times because of their acute cancer diagnoses and the things we talked about. But still, I think we all understood each other very well right from the start", GM6* | *"Yes, definitely the openness. And also, this good atmosphere—wishing well for others, listening to each other, and maybe sharing helpful tips or something like that", HCP2* |
| **Group set-up** |  |
| Safe environment |  |
| *"The space was just very comfortable—everyone was allowed to finish speaking, everyone was acknowledged, and people could respond to each other. The atmosphere was simply very clear. You could open up without fear that anything would be judged or taken away from you. I really thought that was great", GM4* | n/a |
| Small group size |  |
| *"It was a small group, which I really liked. Because in big groups—which I've experienced before—you have so many people and have to share so much, and then there's always a lot of questioning. But that wasn't the case here", GM8* | n/a |
| Heterogenous group constellation |  |
| n/a | *"I wouldn't change the group's makeup—the different levels of anxiety and comorbidities, like Axis II disorders ((personality disorders)) or other anxiety disorders—because that reflects real life. This isn't a controlled observation setting, but a real-world care situation within the healthcare system", HCP1* |

In the first sub-domain “***Exchange with others***”, we identified five mechanisms of change.

*Shared experiences:* Others sharing similar experiences made group members feel relieved and reduced feelings of abnormality. The HCPs noted the group’s effectiveness in making participants feel understood and less alone in their challenges. They highlighted participants’ ability to discuss fears about medical examinations, perceiving a calming effect as participants realized others shared similar fears.

*The act of sharing:* Conversing in the group and talking about problems in itself was found to be helpful. For example, group members could share concerns in light of the SARS-CoV-2 pandemic. For some, taking the space and sharing about themselves was assumed to have the potential to increase their self-efficacy.

*Learning from each other:* Some group members felt that advice from fellow group members provided them with helpful coping strategies and perspectives. HCPs agreed that group members could benefit from each other’s experiences, advice, and support.

*Comparison with others:* Hearing others’ stories puts one’s own situation into perspective. It also reveals when one has not received the best possible care by comparing experiences with other group members. Hearing about good experiences with physicians might also help group members to seek good care experiences as well.

*Humor:* In addition, one group member found it helpful to have a laugh amongst each other about things that they could not laugh about with people without a cancer diagnosis.

In the second sub-domain “***Formation of social connections***”, we identified two mechanisms of change.

*Feeling of community:* Most group members found solace in the camaraderie of the group, fostering social contacts. Some group members even became friends beyond the group sessions.

*Participation in social interactions:* Engaging in social activities within the group helped alleviate social anxieties of some group members.

In the third sub-domain “***Mutual appreciation***”, we identified three mechanisms of change.

*Validation by others and empathetic interactions:* Most group members felt understood and validated when sharing experiences. HCPs observed that group participants treated each other with empathy and without judgment.

*Building trust among group members:* Moreover, some group members and HCPs reported that trust and a sense of belonging were established within the group.

*Open and supportive group atmosphere and group dynamics*: The positive and open atmosphere made most group members feel comfortable. HCPs noted the helpfulness of an open, supportive, and trusting atmosphere with everyone having sufficient space.

In the fourth sub-domain “***Group set-up***”, we identified three mechanisms of change.

*Safe environment*: Some group members felt comfortable to open up in front of the group and express themselves, despite sharing difficult content.

*Small group size:* One group member appreciated the small group size and thus not having to expose themselves in front of too many people.

*Heterogeneous group constellation:* HCPs found it helpful that group members differed in cancer staging, anxiety, comorbidities, age, sociocultural background, and educational level.

#### Mechanisms of change related to the dog

We identified seven mechanisms of change related to the dog. Corresponding illustrative quotes are pro-vided in Table 3.

**Table 3.** Illustrative quotes by the domain Mechanisms of change related to the dog. –.

|  |  |
| --- | --- |
| ***Mechanisms of change related to the dog*** |  |
| Quotes by group members | Quotes by HCPs |
| Catalyzation of group mechanisms |  |
| *"I have to admit, I was always really looking forward to the dog. Even when I wasn't motivated to go, I would still think, well, the dog is there.", GM4* | *"I felt that the dog was a connecting element (...) it helped to create connection and made the group feel together", HCP1* |
| Experiencing joy |  |
| *"Yeah, like I said, just a great dog and really an asset to the group. At least that's how it felt to me. There were several participants who also have dogs themselves, and I think for them it was even more significant. Like I said, I like dogs, but I'm not a huge dog person. For me, it was simply a positive addition because, well, as I said, this animal just radiates something that helps calm you down", GM6* | *"I believe it's fun not only for us HCPs, but also, hopefully, for the participants. It adds something positive, so the group isn't just a place where you talk about the tough parts of your life", HCP3* |
| Feeling safe |  |
| *"I really appreciated having the dog around because it provided a bit of a distraction. Some things are hard to hear or emotionally heavy when others share their experiences, and the dog helped lighten the mood a bit, so it wasn't all negative", GM7* | *"I thought it ((the dog)) was a good anchor for some of the anxious patients. While they were talking about uncomfortable or anxiety-provoking topics, they could distract themselves by petting it or something like that. I really felt that it helped them open up more", HCP1* |
| Building trust with the dog |  |
| *"It was like it really sensed what was happening. Then it'd move on to the next person and seek some affection. So yeah, it just really matched the overall positive atmosphere", GM6* | *"What I think is wonderful is that some people who were initially a bit wary of dogs eventually found the courage to take a photo with it or told their families about the therapy dog, even sharing that they dared to pet it. There's a lot of pride in overcoming that hesitation and allowing the dog to get close or engaging with it in some way", HCP3* |
| Experiencing self-efficacy |  |
| *"Sometimes I felt like I moved, and then the dog would come running over, and I thought ((amused)), 'Oh no, here it comes, what do I do?' And maybe through that I got to understand myself better", GM2* | *"You know, the dog is a big dog, a large Golden Retriever, and I think realizing that it reacts to me, or maybe even does what I say, or lets me be, or that I dare to pet it—I believe that involves overcoming fears, building courage, and feeling like I can achieve something if I set my mind to it. So maybe it's more than just a side effect. Maybe it's actually a positive mechanism of change", HCP2.* |
| Feeling accepted |  |
| *"Yeah, that definitely helped me because the dog is just so calm and treats everyone the same, and I really like that", GM7* | *"It's just there—so carefree and present in the moment. It doesn't pretend, and I think that's something really special about animals; it's such a wonderful addition. (...) with it, it's not really about wondering, 'Does it like me or not?' It's just there—friendly, relaxed, wandering around. And I think that can actually make it easier to connect with it than with people, who might judge you or think badly of you", HCP3* |
| Learning perspective taking |  |
| *"Even now, when I meet dogs in the park or while taking a stroll, I try more to put myself in their place—and I actually benefit from that. I mean, it's hard to describe, but I feel like it helped me become a bit better at empathizing, maybe not just with dogs, but with others in general too", GM2* | n/a |

*Catalyzation of group mechanisms*: The presence of the dog particularly motivated some group members to sign up for the group and made them look forward to the sessions. Other group members had initial reservations towards dogs, but then perceived the dog’s presence as enriching. HCPs also observed facilitation of group interaction per the dog’s presence.

*Experiencing joy*: A positive effect of the dog on mood was reported. Many group members enjoyed padding the dog or playing with it. For group members without prior dog contact, this was a novel and joyful experience. Correspondingly, HCPs noted group members’ enjoyment and mood enhancement.

*Feeling safe:* Group members expressed how the dog’s presence reduces anxiety and fostered a sense of comfort and calm. HCPs noted that interactions with the dog seemed to instill security in group members. Additionally, the dog served as a constructive distraction, redirecting attention away from negative thoughts and emotions.

*Building trust with the dog:* Most group members learned to trust the dog due to its character and being dependable. The dog seemed to sense who was open to him coming close and to what extent. HCPs said that the dog displayed sensitivity by providing comfort to individuals who were upset. Group members and HCPs reported that over the span of the sessions, some group members were able to address their fear of dogs.

*Experiencing self-efficacy*: One group member noticed their effect on the dog (e.g., dog’s behavior after they called him). HCPs assumed that finding the courage to call the dog to them or pet it, could have an effect on self-efficacy.

*Feeling accepted:* One group member felt accepted as the dog displayed an unprejudiced nature. Thus, group members and HCPs agreed that it treated everyone equally and was nonjudgmental.

*Learning perspective taking:* One group member reported that they learned from the dog to empathize better with others and to be more aware of their needs.

#### Mechanisms of change related to the healthcare professionals

We identified three mechanisms of change related to the healthcare professionals. Corresponding illustrative quotes, including those regarding the therapeutic interventions, are provided in Table 4.

**Table 4.** Mechanisms of change related to the HCPs and therapeutic interventions. –.

|  |  |
| --- | --- |
| ***Mechanisms of change related to the healthcare professionals*** |  |
| Quotes by group members | Quotes by HCPs |
| Healthcare professionals' appreciation, empathy and attentiveness |  |
| *"((talking about the HCPs)) Basically, they tell you, "Hey, right now you're the main person or one of the main persons here," and that they care about you and understand you. That's the message there, and I think it's a really meaningful feeling. It's also positive for your self-esteem — experiencing that isn't a bad thing at all", GM1* | n/a |
| Team cohesion among HCPs |  |
| n/a | *"I have a lot of respect for HCP3 ((clinical psychologist)) and know that she's very experienced, so I felt very comfortable working with the co-therapists. HCP2 ((dog handler)), too, is a warm and empathetic person with a lot of experience in therapeutic group settings and with patients. I really felt I was in a strong team of therapists, or maybe three and a half if you count the dog. It gave me the sense of being well supported, with capable people by my side. It was just a really good match", HCP1* |
| Clear structures |  |
| n/a | *"I think what was important from our side was mainly to provide the framework and to moderate the group. So, regularly checking in — is everyone getting a fair chance to speak? Is someone taking up too much space? Is someone maybe too quiet or not coming through enough? Just gently steering things, observing where we are and where we're heading", HCP3* |
| ***Mechanisms of change related to the therapeutic interventions*** |  |
| Gaining knowledge and concrete guidance |  |
| *"It's really about trying to, and in my view that's the main thing that happens there, something I'd call help for self-help. Basically, being given tools to better deal with whatever kind of fears or problems come up, and not letting them take over completely", GM1* | *"And I also think that those classic psychoeducational elements — like the idea that anxiety doesn't just keep rising indefinitely, but eventually subsides — were important for them. As a foundation to help them understand their emotional experiences, I believe that was really valuable", HCP1* |
| Learning relaxation and mindfulness |  |
| *"For me, it was really helpful to do that breathing exercise. I'd say it was the main resource I took away from the group experience. Because I still use it today—several times a week whenever I feel like I need it", GM5* | *"The exercises were mainly attention-focusing techniques that I felt the patients could really relate to. For example, the five, four, three, two, one technique—super simple exercises. I also had the impression that many were able to take these with them, including the introduction to mindfulness-based interventions and relaxation exercises. I do believe that through the experience, they really gained something valuable", HCP1* |

*Healthcare professionals’ appreciation, empathy and attentiveness:* Most group members found the positive and empathetic attention they received from the HCPs helpful, especially for their self-esteem. It made them feel valued and appreciated. Furthermore, some group members valued the clinical psychologists checking in with them by phone during the time sessions were on halt due to the SARS-CoV-2 pandemic.

*Team cohesion among HCPs:* HCPs experienced their collaboration amongst each other as reliable and supportive.

*Clear structures:* HCPs perceived having a dependable structure for each session as important. They also deemed it necessary that they moderated the group members’ contributions by limiting some and encouraging others.

#### Mechanisms of change related to the therapeutic interventions

We identified two mechanisms of change related to the therapeutic interventions.

*Gaining knowledge and concrete guidance:* Group members appreciated receiving information about psychological and related content (i.e., psychoeducation). They also valued guidance on how to deal with specific situations. HCPs added that they used a resource-oriented psychotherapeutic approach and tried to base their guidance on experiences shared by group members.

*Learning relaxation and mindfulness:* Group members and HCPs valued having relaxation and mindfulness exercises incorporated, especially for coping with anxiety and anger. Group members tried integrating these exercises into situations outside of the sessions.

## DISCUSSION

With this exploratory cross-sectional qualitative study, we assessed the mechanisms of change experienced by group members, clinical psychologists, and the dog handler in a DAPOGI for highly anxious cancer survivors. Group members and HCPs reported 24 mechanisms of change, sorted into interactions in the group, the dog, the group set-up, the HCPs, and concrete strategies. There was wide overlap between mechanisms of change perceived by group members and HCPs.

We found substantial overlap between the mechanisms of change in this DAPOGI and existing literature on mechanisms of change of group therapy such as Yalom’s therapeutic factors of group therapy [10]. If these factors were to be applied to the DAPOGI, we suggest that some originally described for human-to-human interactions should be extended to interactions with the dog. By doing so, dog-specific mechanisms of change that were found in our study can also be mapped to Yalom’s therapeutic factors. This applies for example to 1) Yalom’s factor *development of socializing techniques* (i.e., learning and testing new ways to engage with each other) and our dog-specific mechanisms of change *building trust with the dog* and *experiencing self-efficacy*; 2) Yalom’s factor *imitative behaviors* (i.e., learning by observing others) and our dog-specific mechanism of change *learning perspective taking*; and 3) Yalom’s factor *inter-personal learning* (i.e., corrective emotional experiences) and our dog-specific mechanism of change *feeling safe* and *feeling accepted*. Some of the mechanisms of change in the DAPOGI corresponded to more than one of Yalom’s factors. For example, our mechanisms of change *learning from each other* can be mapped to Yalom’s *imparting information* and *imitative behaviors*, and our mechanisms of change *validation by others and empathic interactions* can be mapped to Yalom’s *altruism* and *group cohesiveness*. Only one of Yalom’s therapeutic factors, *corrective recapitulation of the primary family group*, was not found in our study, possibly because the DAPOGI is not a psychodynamic or psychoanalytic intervention. Furthermore, we identified the following additional mechanisms of change in the DAPOGI that did not correspond to any of Yalom’s factors: *humor, supportive group atmosphere, team cohesion among HCPs, clear structures, safe environment, small group size*, and *heterogeneous group constellation*. The majority of these mechanisms of change seem to relate to contextual factors. Further investigations should explore if and how these findings could add to Yalom’s therapeutic factors.

Besides general therapeutic factors of group psychotherapy, the presence of the dog enriched the group intervention at hand, aligning with mechanisms of effectiveness described by Shen et al [18]. The dog’s presence was found to have a profound impact at multiple levels: individual group members reported increased self-confidence and self-efficacy, while interactions among participants were facilitated, fostering camaraderie and group cohesion. Notably, the dog functioned beyond a mere strategy or tool; rather, it assumed the role of an additional group member or co-therapist. These findings align with previous studies [16,17,27], although previous research has primarily emphasized social aspects of the dog’s presence, like initiating conversations or trust building. Although much of the existing literature comes from pediatric settings, our results suggest that these beneficial dynamics extend to adults. Similar mechanisms, such as enhancing confidence, facilitating social interaction, and strengthening group cohesion, also seem to exist in adult populations. This finding highlights the broad applicability of AAIs across age groups. However, our study uncovered additional coping processes induced by AAI, such as gaining perspective-taking skills through interactions with the dog and experiencing the joy the dog brought to the group dynamic. Notably, the presence of the dog in this group psychotherapy context, tailored for highly anxious cancer survivors, appeared to provide additional benefits compared to traditional group therapy settings without animal involvement.

### Strengths and limitations

As little is known about specific mechanisms of change of DAPOGIs, the exploratory and inductive qualitative approach is a major strength of this study. Nevertheless, as for any exploratory research, the results are hypotheses requiring further testing. Furthermore, we analyzed data from different stakeholder perspectives and a heterogeneous sample. Even though purposive sampling and the small sample size pose limitations, we were able to interview group members who varied in age, education level, and cancer staging. Only one run of the DAPOGI could be included, but all invited group members from the DAPOGI agreed to participate in this study. A second limitation was the involvement of PH, as one of the two clinical psychologists in the DAPOGI, as an interview participant in this study, and as the principal investigator of this study. We mitigated this double role by minimizing PH’s role in participant recruitment and data collection and by triangulating perspectives during data analysis. Regarding data analysis, a further limitation was that primarily one researcher undertook the qualitative coding. However, a second researcher gave extensive feedback.

### Clinical and research implications

This study suggests that DAPOGIs could be valuable and effective interventions to support psychologically distressed cancer survivors. Further research is needed to confirm this hypothesis. Next steps include exploratory and confirmatory clinical trials in order to thoroughly develop DAPOGIs as an evidence-based intervention. The findings of this study are an encouraging step toward developing and offering DAPOGIs for cancer survivors as part of interdisciplinary and holistic high-quality cancer care.

## CONCLUSION

The mechanisms of change of the DAPOGI for highly anxious cancer survivors identified in this study were found to exceed those already known from general group psychotherapy. The presence of the dog seems to have increased positive experiences of group members and HCPs and active mechanisms of change. This suggests that incorporating a therapy dog could enhance established psycho-oncological group interventions. Confirmatory research and clinical trials would be valuable next steps towards establishing a nuanced understanding of the effects and mechanisms of change of DAPOGIs.

## Supporting information

Suppl. Material 1 - COREQ

Suppl. Material 2a - Interview guide group members

Suppl. Material 2b - Interview guide HCPs

Suppl. Material 3 - Demographic questionnaire

Suppl. Material 4 - Coding system

## DECLARATIONS

### Ethics Statement

This study was carried out according to the latest version of the Helsinki Declaration of the World Medical Association. The study was approved by the Psychological Ethics Committee of the Center for Psychosocial Medicine of the University Medical Center Hamburg-Eppendorf (LPEK-0232). Standards of research ethics were met. Participants provided written informed consent to participate in this study.

### Author Contributions

CT contributed to this study by conducting data collection, formal analysis and drafting of the first draft as part of her masters’ thesis. MR contributed to the formal analysis of this study and was involved in the writing process of the original manuscript. PH and IS jointly conceptualized the study, devised the research methodology and managed project administration. PH provided supervision throughout the process and was involved in the writing of the manuscript, focusing on reviewing and editing. All authors gave feedback to and consented on the final manuscript.

## Acknowledgments

We thank everyone who supported our study, especially all participating group members. Furthermore, we thank Lisa Brandes for her contributions to this manuscript.

## Data Availability Statement

The data supporting the findings of this study are available from the corresponding author on reasonable request.

## Declaration of AI and AI-assisted technologies in the writing process

During the preparation of this work the authors used the software ChatGPT to improve readability and language. After using this tool, the authors reviewed and edited the content as needed and take full responsibility for the content of the publication.

