## Supplementary material for "Mechanisms of change in a dog-assisted psycho-oncological group intervention: An explorative qualitative interview study": Suppl. Material 2a - Interview guide group members

**SUPPLEMENTARY FILE 2a**

**Interview guide for group members**

**Preparation for Telephone Interview**

- Audio device
- Received informed consent?
- Duration of the interviews: approximately 30 - 60 minutes

Introduction

**Procedure for Individual Interviews**

- INTERVIEWER calls the participant at the agreed-upon time: "Good day, Mrs./Mr. XY, my name is Cheyenne Topf. As you know, I'm calling for our scheduled phone interview. The interview will last approximately 30-60 minutes; I assume the timing still works for you?"
- Note that I unfortunately did not participate in the group therapies due to a possible confusion with the intern.
- At the beginning: "I have received your consent form by mail. Have you already filled out your questionnaire? Do you have any further questions? If not, we can proceed directly to the interview. As discussed, I will now turn on the audio recording device."
- TURN ON AUDIO RECORDING DEVICE!
- Interview goal: "With the interviews, as you know, I want to find out how you experienced the group therapy overall. It covers both the group therapy itself and how it was for you that the therapy was accompanied by a dog."
- "I will now ask you a few questions one by one, and please feel free to say anything that comes to your mind. For the interview, it is important that you feel confident sharing your opinions and experiences openly and honestly. There is no right or wrong here. Feel free to share both critical and positive impressions."

Guiding Questions

To start, I would like to ask you a few general questions:

1. What experiences have you had with dogs before joining this dog group?
2. Have you perhaps had experiences with therapy dogs or other therapy animals elsewhere?
3. Had you already had experiences with psychological groups before the dog group?

Now, let's gradually get more specific about the dog group. Please recall the time when you first learned about the group therapy. This could have been through a therapist or from a flyer.

4. What were your initial thoughts?
5. Why did you want to participate in the group?
6. What expectations did you have for the group therapy?

Now, I'd like you to remember the group sessions. There was an initial introductory session in January 2020, and before the pandemic, there were 3 sessions held in the ground-floor group room. After the break, there were 3 more sessions in the summer of 2020.

7. Do you remember how many sessions you attended?

Please remember how it was to sit together with the other participants, Psychologist 1, Psychologist 2, Dog Handler 1, and Dog 1.

8. What was your first impression when you attended the introductory group session for the first time?
9. How was it to get to know Dog 1?

From now on, we're talking about all group sessions:

10. What did you experience as particularly helpful in the group?
    [Ask multiple times: What else did you find helpful until the participant says that they can't think of anything else.]

i. If not mentioned yet:

What helpful resources were you able to gain through group therapy? (If not directly understood: Resources can also be considered as individual "sources of strength"; these are needed to solve problems or cope with difficulties. These can be, for example, family or friends, but also personal resources such as character traits, values, and experiences, or learned skills and competencies.)

What strategies and exercises did you find helpful? In which problems could the group help you well? e.g., strategies to achieve calmness, mindfulness, or self-compassion, strategies in dealing with fears or in reducing anxiety, increasing self-efficacy and the sense of coherence (acceptance of the situation), universality of suffering (feeling not alone with problems)

What did you experience as less helpful?

[Ask multiple times: What else did you find less helpful until the participant says that nothing else comes to mind.]

i. If not mentioned yet:

Which strategies and exercises did you find less helpful? In which problems could the group help you less effectively?

11. Did anything potentially bother you about the group sessions?
    i. [If yes:] What was it?

I would like to specifically discuss the therapy dog, Dog 1.

12. How did you experience his presence in the group?

a. What was nice/pleasant/helpful?

i. If not mentioned yet:

How did Dog 1's presence influence your well-being? What impact did Dog 1 have on your anxiety?

b. What was less helpful/unpleasant?

13. You mentioned some points above that were less helpful. What improvement ideas do you have for the group with a therapy dog?
    i. [If experiences with group therapies are reported]:

You mentioned that you have participated in the (specific) group. How do your experiences in different groups differ?

Summary

1. When you reflect on everything we have discussed today, which aspects are particularly important to you?
2. Is there something we haven't discussed yet that should definitely not be missed?

Ending

Acknowledgment and farewell

End audio recording
