## Supplementary material for "Mechanisms of change in a dog-assisted psycho-oncological group intervention: An explorative qualitative interview study": Suppl. Material 2b - Interview guide HCPs

**SUPPLEMENTARY FILE 2b**

**Interview guide for clinical psychologists and dog handler**

**Preparation for Telephone Interview**

- Audio recording device
- Received informed consent?
- Duration of the interviews: approximately 30 - 60 minutes

**Procedure for Individual Interviews**

Introduction

- INTERVIEWER calls the participant at the agreed-upon time: "Good day, Mrs. XY, my name is Cheyenne Topf. As you know, I'm calling you as part of our scheduled telephone interview. The interview will last approximately 30-60 minutes. I assume the timing is still suitable for you?"
- At the beginning: "I have already received your consent form. Do you have any further questions? If not, we would like to start the interview. As discussed, I will now turn on the audio recording device."
- TURN ON AUDIO RECORDING DEVICE!
- Purpose of the interview: "With the interviews, I want - as you know - to find out how you, as a therapist, experienced the group therapy overall. It involves both the group therapy itself and how it was for you that the therapy was accompanied by a dog."
- "I will now gradually ask you some questions and simply ask you to say everything that comes to mind. For the interview, it is important that you feel free to describe your opinions and experiences openly and honestly. There is no right or wrong here. Feel free to share both critical and positive impressions."

Guiding Questions

To start, I would like to ask you a few general questions:

1. What experiences have you had with dogs before this dog group?
2. What previous experiences have you had with therapy dogs or other therapy animals before this dog group?
3. What previous experiences have you had with conducting psychological groups before this dog group?
4. What prior experiences in treating oncological patients had you gained before this dog group? If yes and not mentioned yet:
a. How many years of experience do you have in treating oncological patients?

Now I would like to gradually delve into the dog group.

5. What expectations did you have in advance of the group therapy?

Now, please recall the group sessions. There was an introductory session in January 2020, and then, before the pandemic, there were 3 sessions in the ground-floor group room. After the break, there were another 3 sessions in the summer of 2020. Recall how it was to sit together with the participants, other therapists, and Dog 1.

6. What was your initial impression when you attended the introductory group session for the first time?

**(Not applicable to Dog Handler 1)**

7. How was it to get to know Dog 1?

**(Applicable to all)**

8. How do you think it was for the patients to get to know Dog 1?

From now on, we'll focus on all group sessions:

9. What did you personally find particularly helpful?

[Ask multiple times: What else did you find helpful until the participant has nothing more to add.]

[Ask what patients found helpful if not mentioned.]

i. If not mentioned:

What strategies and exercises did you find helpful? To the best of your judgment, in which problems could the group be of assistance?

10. What did you personally find less helpful/challenging?

[Ask multiple times: What else did you find less helpful until the participant has nothing more to add.]

[Ask what patients did not find helpful if not mentioned.]

i. If not mentioned:

What strategies and exercises did you find less helpful? To the best of your judgment, in which problems could the group be less helpful?

11. Was there perhaps something that bothered you in the group sessions?
    i. [If yes:] What was it?

I would like to specifically discuss therapy dog 1 again:

12. How did you experience his presence in the group?

[Ask what patients found helpful if not mentioned]

a. What was, in your opinion, nice/pleasant/helpful?

i. If not mentioned:

To the best of your judgment, what influence did the presence of dog 1 have on well-being?

To the best of your judgment, what influence did dog 1 have on fears?)

b. What was, in your opinion, less helpful/unpleasant?

13. You mentioned a few points above that were less helpful. What ideas for improvement do you have for the future implementation of a group with a therapy dog?
    i. [If experiences with group therapies are reported]:

You mentioned that you already have experience in treating oncological patients as well as conducting group therapies. How do your experiences differ in treatment, both in individual therapies and in different group settings?

Summary

1. When you reflect on everything we've discussed today, which aspects are particularly important to you?
2. Is there anything we haven't discussed so far that should not be overlooked?

Ending

Acknowledgment and farewell

End audio recording
