## Supplementary material for "Mechanisms of change in a dog-assisted psycho-oncological group intervention: An explorative qualitative interview study": Suppl. Material 3 - Demographic questionnaire

**SUPPLEMENTARY FILE 3**

**Evaluation of the Dog-Assisted Psycho-Oncological Group Therapy: Questionnaire for Participants**

**Dear participants,**

**with this questionnaire, we would like to gather some anonymous information about you and your experience with psychological services and with dogs. Most questions can be answered by checking the appropriate boxes. For other questions, you can enter your response in the open text fields. It should take approximately 5 minutes to complete the questionnaire.**

**We will not link this information with the data from the telephone interview, but will only use it for the description of the group. For this reason, please do not provide a name on the questionnaire. The data will be analyzed at the Department of Medical Psychology of the University Medical Center Hamburg-Eppendorf by Master's student Cheyenne Topf.**

**We are grateful for your participation in this project and thank you warmly for your support.**

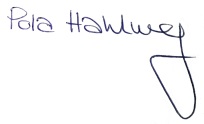

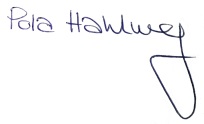

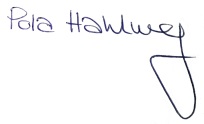

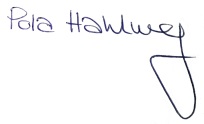

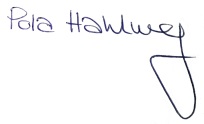

Dr. Pola Hahlweg	Cheyenne Topf

(Principal investigator)	(Master’s student)

| **Personal information**  Please provide some information about yourself: | | | | | | | | | | | | | | | |
| --- | --- | --- | --- | --- | --- | --- | --- | --- | --- | --- | --- | --- | --- | --- | --- |
| 1. | **Today’s Date** | ______________________ | | | | | | | | | | | | | |
| 2. | **Age** | ________ (in years) | | | | | | | | | | | | | |
| 3. | **Gender** | 🞎 | Female | 🞎 | Male | | | 🞎 | Other/no response | | | | | | |
| 4. | **Highest (Educational) Degree** | 🞎 | No formal education | 🞎 | Lower Secondary Education | | | 🞎 | Intermediate Secondary Education | | | 🞎 | | High School Diploma/General Certificate of Education | |
|  |  | 🞎 | Technical/Vocational School Diploma | 🞎 | College/University Degree | | | 🞎 | Other (please specify):  _____________________________________ | | | | | | |
| 5. | **Current work situation?** | 🞎 | Employed  🞎 Full-time  🞎 Part-time | 🞎 | Homemaker | | | 🞎 | Retired  🞎 Due to age  🞎 Disability pension  🞎 Early retirement, but no disability pension | | | | | | |
|  |  | 🞎 | Unemployed | 🞎 | Other (please specify):  __________________________________________________ | | | | | | | | | | |
| 6. | **Are you currently on sick leave?** | | | 🞎 | No | | | 🞎 | Yes | | | | | | |
| 7. | **Do you have dogs living in your household?** | | | 🞎 | Yes, currently | | | 🞎 | Yes, in the past | | | | 🞎 | | No |
| **Information about your illness and experiences with psychosocial services.** | | | | | | | | | | | | | | | |
| 8. | **Which cancer type have you been diagnosed with?**  **___________________________________________________________________________________________________** | | | | | | | | | | | | | | |
| 9. | **When was your initial diagnosis?** (Date of the diagnosis): **___________________________________________________________________________________________________** | | | | | | | | | | | | | | |
| 10. | **Is your condition currently localized or metastasized?** | | | | | 🞎 | Localized | | | 🞎 | Metastasized | | | | |
| 11. \| **Do you have experience with the following support services so far?** Please check all applicable answers: \| \|; \| \| **In the past** \| **In the present**; \| 1) Individual psychotherapy \| 1) 🞎 \| 1) 🞎; \| 2) Group psychotherapy \| 2) 🞎 \| 2) 🞎; \| 3) Psychosocial counseling \| 3) 🞎 \| 3) 🞎; \| 4) Social services \| 4) 🞎 \| 4) 🞎; \| 5) Health care chaplaincy \| 5) 🞎 \| 5) 🞎; \| 6) Self-help groups \| 6) 🞎 \| 6) 🞎; \| 7) Animal-assisted therapy \| 7) 🞎 \| 7) 🞎; \| 8) Other (please specify): **___________________________** \| 8) 🞎 \| 8) 🞎 | | | | | | | | | | | | | | | |

**Thank you!**
